# Stage-specific overall survival with complete five-year follow-up after D2 gastrectomy for gastric cancer at a referral hospital in Shanghai, China: a reference for comparing hospitals

**DOI:** 10.64898/2026.10.12.26365370

**Authors:** Birendra Kumar Sah, Danping Gu, Jian Li, Erzhen Chen

## Abstract

**Background:** Comparing long-term outcomes of gastric cancer surgery between hospitals requires a common reference. Risk-adjusted benchmarks depend on the model and population used to build them and are difficult for another hospital to reproduce. TNM stage is a prognostic classification already recorded for every resected gastric cancer. We report unadjusted stage-specific overall survival from a consecutive cohort with complete follow-up as reference data and propose a minimal dataset for producing comparable figures elsewhere.

**Methods:** The cohort comprised 690 consecutive eligible patients who underwent curative-intent D2 gastrectomy for stage I to III adenocarcinoma at Ruijin Hospital, Shanghai, in 2019. All operations were performed by eight experienced, high-volume consultant surgeons. Stage followed the AJCC 8th edition: pTNM after upfront surgery (n = 570) and ypTNM after neoadjuvant chemotherapy (n = 120). Overall survival was estimated by the Kaplan-Meier method without adjustment.

**Results:** There were 199 deaths, 181 within five years. Every survivor was followed for at least 68.1 months. After upfront surgery, five-year overall survival was 95.0% (95% CI 91.4 to 97.1) for stage I, 83.8% (76.5 to 89.0) for stage II and 56.5% (49.2 to 63.1) for stage III, and was 95.0% in IA and 95.2% in IB, falling to 26.7% in IIIC. After neoadjuvant chemotherapy, five-year survival was 78.6%, 61.9% and 31.3% for ypStage I, II and III. Pooling pTNM and ypTNM lowered stage III five-year survival to 50.2%. The proposed minimal dataset has 16 items, 13 of them recorded during the admission for gastrectomy.

**Conclusions:** These figures are intended as a reference for comparing hospitals rather than as a standard of care. We propose 16 items, most of them routinely recorded, from which another hospital can produce the same table and compare its results by stage.

## Introduction

Gastric cancer ranks fifth worldwide in both incidence and mortality, with more than 968,000 new cases and close to 660,000 deaths in 2022 [1]. Population-based five-year survival differs several-fold between countries [2] and, although it has improved in China, remains below that in Japan and Korea [2, 3]. Outcomes also vary between hospitals within a country, which has prompted volume-based referral and national surgical audits [4-6]. Hospitals, insurers and regulators increasingly compare the results of cancer surgery [7, 8]. For gastric cancer the principal long-term outcome is overall survival, and its strongest determinant is pathological stage [9, 10], so a fair comparison between hospitals has to be made within stage.

The usual approach is risk adjustment, in which a model fitted to a reference population gives each hospital an expected number of deaths and observed deaths are divided by expected. The resulting ratio depends on the covariates in the model, the population used to fit it and the completeness with which each hospital records them [11, 12]. When the model is fitted to the group being audited, the expected total matches the observed total by construction, so the ratios can only rank hospitals against one another. Ranking hospitals on such ratios can also attribute differences in data quality or case-mix to differences in care [7]. Funnel plots show each hospital against a reference with limits for chance [13], but the reference itself still has to be chosen. A benchmark derived in one population also reflects that population’s background mortality, which limits its use in another [14]. In a recent global cohort, case-mix and treatment explained little of the between-hospital variation in most short-term gastrectomy outcomes; the exceptions were 30-day mortality and negative resection margins [8]. International benchmarks for gastrectomy cover short-term outcomes such as morbidity, node yield and 90-day mortality, but not long-term survival [15].

TNM stage does not depend on a fitted model or a reference population. It is recorded for every resected gastric cancer, defined by an international standard and verified through pathology, and it is strongly prognostic [9, 10]. The 8th edition has been validated in large Western datasets [16, 17], and separate ypTNM groups were introduced for patients treated before surgery [18]. Stage-specific survival has been published from national registries and single centres [19-21], but most series have incomplete follow-up, mix staging editions or treatment eras, or combine patients staged before and after neoadjuvant treatment. The Japanese national registry, for example, followed 89.5% of patients registered in 2010 to 2013 for five years [20]. Few report the patient and death counts that another hospital would need for a direct comparison.

We therefore report unadjusted stage-specific overall survival from a consecutive cohort in which every survivor was followed beyond five years, with pTNM and ypTNM reported separately, and propose a minimal dataset from which other hospitals can produce equivalent figures.

## Methods

### Cohort

Of 780 records of gastric resection at the Gastrointestinal Surgery Unit of Ruijin Hospital between 1 January and 31 December 2019, 690 met the eligibility criteria of curative-intent gastrectomy with D2 lymphadenectomy [22] for gastric adenocarcinoma without distant metastasis. Ninety were excluded for a non-adenocarcinoma or other ineligible diagnosis (34), stage IV disease (50), pathological complete response (4) or a duplicate record (2); no exclusion was based on outcome. Operations were performed by eight consultant surgeons, each with established experience in gastric cancer surgery, who performed between 43 and 139 of the included operations. Resection was R0 in 678 patients and R1 in 12.

### Staging

Stage was assigned from the final pathology report according to the AJCC 8th edition [9]. CT images and pathology slides were not re-reviewed; all data were taken from the original reports as recorded in routine practice, so that the figures reflect real-world reporting and can inform the design of future datasets. Clinical stage (cTNM) was not used, because in this retrospective cohort the preoperative CT report gave both a T and an N category in only 195 of 690 patients (28.3%), and using pathological stage alone avoids confusion between clinical and pathological stage. Patients who had upfront surgery were staged by pTNM with substages IA to IIIC; those who received neoadjuvant chemotherapy were staged by ypTNM, which uses separate stage groups without substages [18]. Stage groups and substages were derived from pT and pN (all patients were M0) using the corresponding table, with pN3 subdivided into N3a and N3b for substage assignment. The pN category was checked against the number of positive nodes, and discrepancies were resolved by review of the pathology report. In the study database, 18 patients who had upfront surgery and carcinoma in situ or no residual tumour after prior endoscopic resection are recorded as stage I, although the AJCC 8th edition would classify Tis as stage 0 and assign no stage to T0, and they have no substage. They were retained as recorded; excluding them changes stage I five-year survival after upfront surgery from 95.0% to 95.1%.

### Follow-up and endpoint

The endpoint was overall survival from the date of surgery to death from any cause, with survivors censored at last contact. Patients were scheduled for review every three to six months for two years and annually thereafter. For patients who did not attend, vital status was confirmed by linkage to civil registration and death records. The survival cut-off date was 28 July 2025. Survival time was calculated in days and converted to months by dividing by 30. Sixty months (1,800 days) is 26 days short of five calendar years; no death occurred in that interval, so five-year estimates are unaffected.

### Statistical analysis

Survival was estimated by the Kaplan-Meier method [23] for upfront surgery by stage group and by substage, for neoadjuvant chemotherapy by ypStage group, and for all 690 patients by stage group, combining pTNM and ypTNM, because many hospitals report survival this way and the pooled figures show how combining the two changes the estimates. For each stratum we report the number of patients, total deaths, deaths within 60 months, overall survival at one, three and five years with 95% confidence intervals on the log-log scale, and median survival where reached. Strata were compared with the log-rank test. Median follow-up was estimated by the reverse Kaplan-Meier method [24], and completeness of follow-up by the minimum follow-up of living patients. No adjustment was made for patient or treatment factors. Analyses used Python 3.13 with lifelines 0.30. Reporting follows the STROBE statement [25].

### Minimal dataset

We listed the fewest routinely recorded items needed to reproduce the survival estimates in this report and to support routine outcome audit, selecting items that define the population, the treating surgeon, the stage, major morbidity or the survival endpoint. In the dataset, complications are defined by the international consensus list for gastrectomy [26] and graded by the Clavien-Dindo classification [27]. The dataset is intended for routine audit and complements core outcome sets designed for surgical trials [28].

### Supplementary practice data

To describe routine practice in 2019, we tabulated pathology and tissue biomarker findings, and postoperative surveillance and detected recurrence (Supplementary Tables S1 and S2). These data are descriptive, and no survival analysis by these factors was performed. Surveillance data were available for the 534 patients followed at the treating hospital.

## Results

### Cohort and follow-up

Of the 690 patients, 570 had upfront surgery and 120 received neoadjuvant chemotherapy (Table 1). Patients given neoadjuvant chemotherapy more often had total gastrectomy and open surgery, and 53.3% had ypStage III disease. Median follow-up was 74.2 months, and all survivors had at least 68.1 months of follow-up, so five-year survival in each stratum equals the proportion alive at five years. Of 199 deaths, 181 occurred within 60 months.

**Table 1.** Patient and treatment characteristics.

| Characteristic | Upfront surgery<br>(n = 570) | Neoadjuvant<br>chemotherapy (n = 120) | All (n = 690) |
| --- | --- | --- | --- |
| Age, years, median (IQR) | 63 (56 to 70) | 62.5 (54 to 69) | 63 (56 to 69) |
| Age 70 years or older | 144 (25.3) | 28 (23.3) | 172 (24.9) |
| Male | 376 (66.0) | 93 (77.5) | 469 (68.0) |
| Distal gastrectomy | 383 (67.2) | 42 (35.0) | 425 (61.6) |
| Proximal gastrectomy | 13 (2.3) | 0 (0.0) | 13 (1.9) |
| Total gastrectomy | 174 (30.5) | 78 (65.0) | 252 (36.5) |
| Laparoscopic approach | 309 (54.2) | 26 (21.7) | 335 (48.6) |
| R0 resection | 562 (98.6) | 116 (96.7) | 678 (98.3) |
| Nodes examined, median (IQR) | 29 (22 to 39) | 31.5 (23 to 43) | 29 (22 to 39) |
| 16 or more nodes examined | 525 (92.1) | 114 (95.0) | 639 (92.6) |
| Stage I | 241 (42.3) | 14 (11.7) | 255 (37.0) |
| Stage II | 136 (23.9) | 42 (35.0) | 178 (25.8) |
| Stage III | 193 (33.9) | 64 (53.3) | 257 (37.2) |
| Adjuvant chemotherapy, stage I | 45 (18.7) | 10 (71.4) | 55 (21.6) |
| Adjuvant chemotherapy, stage II | 87 (64.0) | 35 (83.3) | 122 (68.5) |
| Adjuvant chemotherapy, stage III | 127 (65.8) | 56 (87.5) | 183 (71.2) |

Values are n (%) unless stated; adjuvant chemotherapy percentages are of patients in that stage. Stage is pTNM after upfront surgery and ypTNM after neoadjuvant chemotherapy. IQR, interquartile range.

### Survival by stage

After upfront surgery, five-year overall survival was 95.0% for stage I, 83.8% for stage II and 56.5% for stage III (Table 2, Figure 1A; log-rank P < 0.001). Five-year survival declined with each substage from IIA to IIIC and was similar in IA and IB (Figure 1B). After neoadjuvant chemotherapy, survival differed between ypStage groups (Figure 1C; log-rank P < 0.001) and was lower at each ypStage than at the corresponding pStage. Pooling the two groups lowered five-year survival from 83.8% to 78.7% in stage II and from 56.5% to 50.2% in stage III.

**Table 2.** Overall survival by stage.

| Stage | n | Deaths,<br>total | Deaths<br>within 60<br>months | 1-year OS, % (95%<br>CI) | 3-year OS, % (95%<br>CI) | 5-year OS, % (95%<br>CI) | Median OS,<br>months |
| --- | --- | --- | --- | --- | --- | --- | --- |
| **Upfront surgery, pTNM** |  |  |  |  |  |  |  |
| I | 241 | 15 | 12 | 99.6 (97.1 to 99.9) | 96.7 (93.5 to 98.3) | 95.0 (91.4 to 97.1) | Not reached |
| II | 136 | 24 | 22 | 98.5 (94.2 to 99.6) | 89.0 (82.4 to 93.2) | 83.8 (76.5 to 89.0) | Not reached |
| III | 193 | 90 | 84 | 95.9 (91.9 to 97.9) | 72.5 (65.7 to 78.3) | 56.5 (49.2 to 63.1) | Not reached |
| **Upfront surgery, pTNM substage** |  |  |  |  |  |  |  |
| IA | 160 | 9 | 8 | 99.4 (95.6 to 99.9) | 96.9 (92.7 to 98.7) | 95.0 (90.3 to 97.5) | Not reached |
| IB | 63 | 5 | 3 | 100.0 | 96.8 (87.9 to 99.2) | 95.2 (86.0 to 98.4) | Not reached |
| IIA | 57 | 8 | 7 | 98.2 (88.2 to 99.8) | 94.7 (84.6 to 98.3) | 87.7 (76.0 to 93.9) | Not reached |
| IIB | 79 | 16 | 15 | 98.7 (91.4 to 99.8) | 84.8 (74.8 to 91.1) | 81.0 (70.5 to 88.1) | Not reached |
| IIIA | 87 | 29 | 26 | 97.7 (91.1 to 99.4) | 78.2 (67.9 to 85.5) | 70.1 (59.3 to 78.6) | Not reached |
| IIIB | 76 | 38 | 36 | 94.7 (86.6 to 98.0) | 73.7 (62.2 to 82.1) | 52.6 (40.9 to 63.1) | 62.2 |
| IIIC | 30 | 23 | 22 | 93.3 (75.9 to 98.3) | 53.3 (34.3 to 69.1) | 26.7 (12.6 to 43.0) | 36.9 |
| **Neoadjuvant chemotherapy, ypTNM** |  |  |  |  |  |  |  |
| I | 14 | 4 | 3 | 100.0 | 92.9 (59.1 to 99.0) | 78.6 (47.2 to 92.5) | Not reached |
| II | 42 | 19 | 16 | 95.2 (82.3 to 98.8) | 73.8 (57.7 to 84.6) | 61.9 (45.5 to 74.6) | Not reached |
| III | 64 | 47 | 44 | 85.9 (74.7 to 92.4) | 37.5 (25.8 to 49.1) | 31.3 (20.4 to 42.7) | 27.5 |
| **All patients, pTNM and ypTNM pooled** |  |  |  |  |  |  |  |
| I | 255 | 19 | 15 | 99.6 (97.2 to 99.9) | 96.5 (93.3 to 98.1) | 94.1 (90.4 to 96.4) | Not reached |
| II | 178 | 43 | 38 | 97.8 (94.1 to 99.2) | 85.4 (79.3 to 89.8) | 78.7 (71.9 to 84.0) | Not reached |
| III | 257 | 137 | 128 | 93.4 (89.6 to 95.8) | 63.8 (57.6 to 69.4) | 50.2 (43.9 to 56.1) | 60.9 |

OS, overall survival; CI, confidence interval (log-log). All survivors were followed beyond 60 months. Substages exclude 18 stage I patients with carcinoma in situ or no residual tumour (Methods). No interval is given where no death occurred. ypStage excludes pathological complete response (ypT0N0), which was outside the cohort definition.

**Figure 1.**
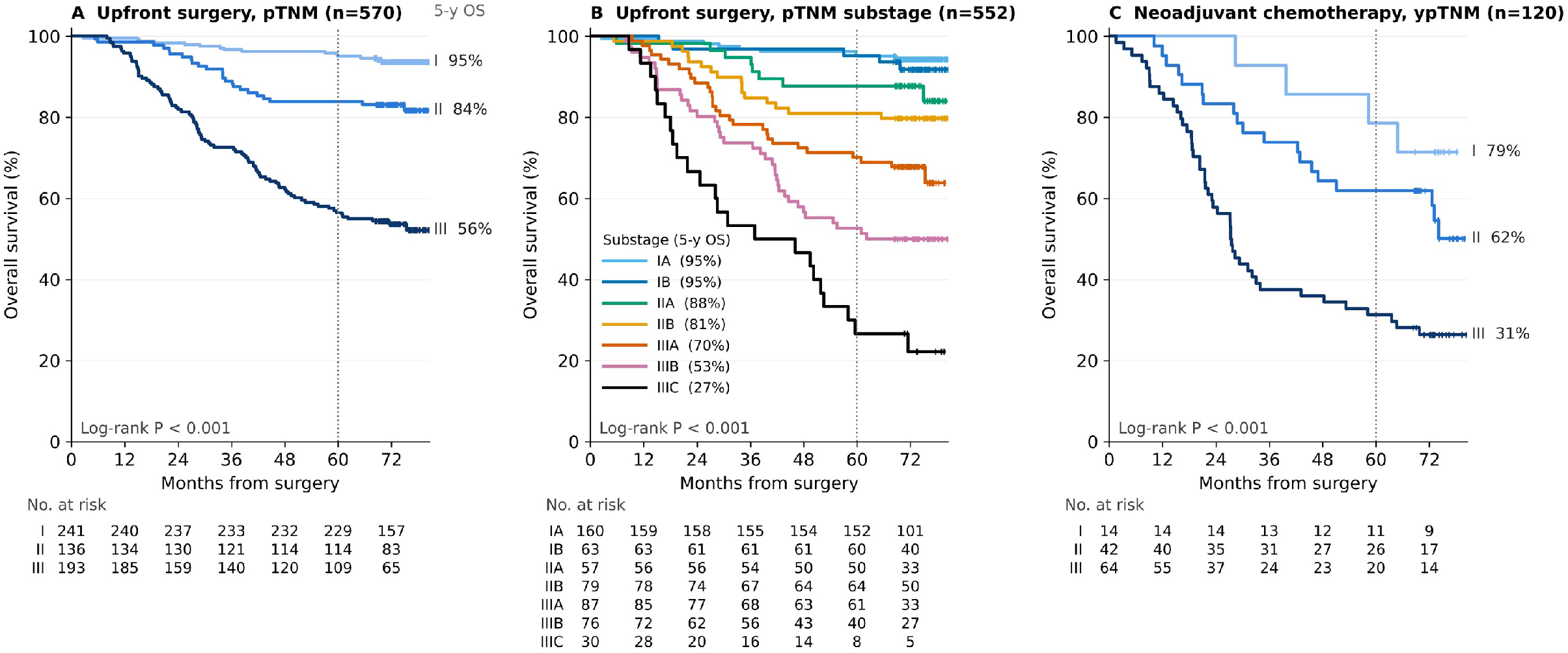
Overall survival by stage. Kaplan-Meier estimates for (A) upfront surgery by pTNM stage (n = 570), (B) upfront surgery by pTNM substage (n = 552; 18 patients with carcinoma in situ or no residual tumour after endoscopic resection are counted in stage I in panel A but have no substage) and (C) neoadjuvant chemotherapy by ypTNM stage (n = 120). Percentages beside the curves, or in the legend of panel B, are five-year overall survival, rounded to whole numbers. Tick marks show censored patients and the dotted line marks 60 months. Numbers at risk are shown below each panel. P values are from the log-rank test.

### Practice in 2019

HER2, mismatch repair and Epstein-Barr virus status were recorded for 82.2%, 95.8% and 92.5% of patients respectively, and PD-L1 for 2.3% (Supplementary Table S1). Among the 534 patients followed at the treating hospital, median hospital-based follow-up was 21.2 months, and 166 of 398 five-year survivors (41.7%) were still seen there at 48 months or later (Supplementary Table S2).

### Minimal dataset

The proposed minimal dataset has 16 items (Table 3). Thirteen are recorded during the admission for gastrectomy. Major morbidity is recorded at discharge and again at 90 days, and vital status and the date of death or last contact require longer follow-up.

**Table 3.** Proposed minimal dataset for stage-specific comparison of survival after gastrectomy.

| No. | Item | Definition or coding | Source |
| --- | --- | --- | --- |
| 1 | Patient identifier | Pseudonymised, unique within the hospital | Hospital information system |
| 2 | Hospital | Coded identifier of the treating hospital | Hospital information system |
| 3 | Age at surgery | Completed years | Hospital information system |
| 4 | Sex | Male or female | Hospital information system |
| 5 | Date of surgery | Day, month, year | Operation note |
| 6 | Operating consultant | Pseudonymised identifier, unique within the hospital | Operation note |
| 7 | Neoadjuvant therapy | None, chemotherapy or other | Treatment record |
| 8 | Extent of gastrectomy | Distal, proximal, total or other | Operation note |
| 9 | Resection status | R0, R1 or R2 | Pathology report |
| 10 | pT or ypT | AJCC 8th edition category | Pathology report |
| 11 | pN or ypN, with nodes examined and nodes positive | AJCC 8th edition category and two counts | Pathology report |
| 12 | pM | M0 or M1 | Pathology report |
| 13 | Stage group | AJCC 8th edition, pTNM substage or ypTNM group | Derived from items 10 to 12 |
| 14 | Major postoperative morbidity | Highest Clavien-Dindo grade [27] at discharge and at 90 days after surgery [26]; grade IIIa or higher is major | Discharge summary or complication record |
| 15 | Vital status | Alive or dead at last contact | Follow-up record or death registry |
| 16 | Date of death or last contact | Day, month, year | Follow-up record or death registry |

## Discussion

This cohort provides stage-specific survival after curative gastrectomy with no patient lost to follow-up before five years, reported with the counts behind each estimate and with pTNM and ypTNM analysed separately. Hospitals that record the items in Table 3 can compare their own results with these by stage.

### Choice of unadjusted stage-specific survival

We did not adjust for factors other than stage. A risk-adjusted ratio cannot be reproduced without the original model and data, and its expected values change with the covariates chosen [11, 12]. A stage-specific survival table needs only the stage, the date of surgery, vital status and the date of death or last contact; the remaining items in Table 3 describe the population, the surgeon and major morbidity for routine audit. Patients within a stage still differ in age, comorbidity, histology and treatment. If those differences are judged too large for fair comparison, it would be better to refine the staging system through the international process that maintains it [9, 10] than for each audit to build its own model. In the meantime, we consider TNM stage the most transparent common basis for comparison between hospitals.

### Interpretation of the reference figures

The figures describe what experienced, high-volume surgeons achieved at one referral centre in one year. They are not intended as a target or as a minimum acceptable level. A difference from this reference, in either direction, does not by itself establish a difference in quality of care. Benchmarks for short-term outcomes after gastrectomy are already available [15], and these figures offer reference data for long-term survival. Surgeons, hospital managers, insurers and regulators can use the same figures and method when discussing differences between hospitals.

A reference has to be interpreted in the setting in which it was produced. Stage-specific survival differs between countries even after adjustment for known prognostic factors; in a comparison of one US and one Korean institution, disease-specific survival after R0 gastrectomy was higher in Korea [29]. The figures reported here reflect a period in which D2 lymphadenectomy [30] and adjuvant chemotherapy after D2 gastrectomy [31, 32] were standard practice, the number of nodes examined (median 29), which affects stage assignment and therefore stage-specific survival [33, 34], and patient selection at a referral centre. Background mortality also differs between populations, and part of any difference in survival between hospitals or regions may reflect deaths unrelated to cancer [14].

### pTNM and ypTNM

Lower survival at each ypStage is expected, because ypTNM describes residual disease after treatment in tumours that were usually more advanced at diagnosis [18]. Perioperative chemotherapy is supported by randomised trials [35-38], but its use varies between hospitals. In this cohort, pooling pTNM and ypTNM lowered stage III survival by 6.3 percentage points, so pooled stage-specific figures depend partly on the proportion of patients treated before surgery. pTNM and ypTNM results should therefore be reported separately, together with that proportion.

### Limitations

This is a single-centre cohort from one year, with data collected in routine practice. Some strata are small, particularly IIIC (30 patients) and ypStage I (14 patients), and their confidence intervals are wide. Deaths from all causes were included, cause of death was not analysed, recurrence was described but not analysed as an outcome, and comorbidity was not recorded in a standard form. Patients with pathological complete response were outside the cohort, so ypStage figures apply to patients with residual tumour. Follow-up spanned the COVID-19 pandemic, which may have added deaths across all stages.

### Implications

Most of the 16 items are recorded routinely by hospitals that resect gastric cancer; the 90-day complication check and follow-up of vital status require added effort. In this cohort, vital status was complete, although fewer than half of five-year survivors followed at Ruijin Hospital were still seen there at four years or later, so survival ascertainment cannot rely on clinic visits alone. For each stage, a hospital should report the number of patients, deaths within 60 months, five-year survival with its confidence interval and the number censored before 60 months, which shows whether the estimate rests on complete follow-up. Reporting the proportion of patients with 16 or more nodes examined helps to judge stage migration. Results from several hospitals can then be shown against the reference in a funnel plot, which allows for the play of chance at each hospital’s volume [13]. We propose these items as the basis for a common minimal dataset, starting with hospitals in Shanghai. If more hospitals report in this format, a pooled multicentre reference could replace the single-centre one presented here.

## Conclusions

In a consecutive cohort with complete five-year follow-up, unadjusted five-year survival after curative gastrectomy fell from 95.0% in pStage I to 56.5% in pStage III and was lower at each ypStage after neoadjuvant chemotherapy. These figures can serve as a reference for comparing overall survival after gastrectomy between hospitals by stage, and the proposed 16-item dataset is designed to let other hospitals produce equivalent figures.

## Supporting information

Supplementary Tables S1 and S2: pathology and tissue biomarker testing, and follow-up, surveillance testing and detected recurrence

## Declarations

### Ethics approval and consent

The study was approved by the Ruijin Hospital Ethics Committee (approval no. 2025-594, 18 September 2025), which waived individual consent for this retrospective analysis of de-identified data.

### Data availability

Table 2 gives the number of patients and deaths behind each estimate, and Figure 1 gives the numbers at risk. The de-identified patient-level dataset may be made available on reasonable request, subject to institutional approval. Analysis code that reproduces all tables, the figure and the numbers derived from the data is available from the corresponding author.

### Author contributions

B.K.S.: conceptualisation, methodology, formal analysis, data curation, visualisation, writing (original draft). D.G.: data curation, validation, writing (review and editing). J.L.: validation of the survival analyses, project administration including ethics approval, writing (review and editing). E.C.: supervision, resources, critical revision of the manuscript. B.K.S. and D.G. contributed equally as co-first authors; B.K.S. and E.C. jointly supervised the work. All authors approved the final manuscript.

