## Supplementary Tables S1 and S2: pathology and tissue biomarker testing, and follow-up, surveillance testing and detected recurrence for "Stage-specific overall survival with complete five-year follow-up after D2 gastrectomy for gastric cancer at a referral hospital in Shanghai, China: a reference for comparing hospitals"

These tables describe routine practice at Ruijin Hospital for the 690 patients operated on in 2019, covering pathology and tissue biomarker findings, and postoperative follow-up and detected recurrence. They are descriptive and are provided so that other hospitals can compare their own practice with that of this centre in 2019. No survival analysis by these factors is presented.

### Table S1. Pathology and tissue biomarker testing by stage group, 2019 (n = 690)

| **Characteristic** | **Upfront, stage I (n = 241)** | **Upfront, stage II (n = 136)** | **Upfront, stage III (n = 193)** | **Neoadjuvant, ypStage I to III (n = 120)** | **All (n = 690)** |
|---|---|---|---|---|---|
| **Differentiation** | | | | | |
| G1 | 16 (6.6) | 0 (0.0) | 0 (0.0) | 0 (0.0) | 16 (2.3) |
| G1 to G2 | 22 (9.1) | 0 (0.0) | 0 (0.0) | 2 (1.7) | 24 (3.5) |
| G2 | 48 (19.9) | 30 (22.1) | 12 (6.2) | 15 (12.5) | 105 (15.2) |
| G2 to G3 | 67 (27.8) | 58 (42.6) | 76 (39.4) | 41 (34.2) | 242 (35.1) |
| G3 | 67 (27.8) | 45 (33.1) | 103 (53.4) | 58 (48.3) | 273 (39.6) |
| Not recorded | 21 (8.7) | 3 (2.2) | 2 (1.0) | 4 (3.3) | 30 (4.3) |
| **Signet-ring cell component** | | | | | |
| Yes | 28 (11.6) | 8 (5.9) | 9 (4.7) | 11 (9.2) | 56 (8.1) |
| No | 212 (88.0) | 128 (94.1) | 184 (95.3) | 109 (90.8) | 633 (91.7) |
| Not recorded | 1 (0.4) | 0 (0.0) | 0 (0.0) | 0 (0.0) | 1 (0.1) |
| **Lymphovascular invasion** | | | | | |
| Yes | 23 (9.5) | 57 (41.9) | 166 (86.0) | 61 (50.8) | 307 (44.5) |
| No | 214 (88.8) | 74 (54.4) | 24 (12.4) | 57 (47.5) | 369 (53.5) |
| Not recorded | 4 (1.7) | 5 (3.7) | 3 (1.6) | 2 (1.7) | 14 (2.0) |
| **Perineural invasion** | | | | | |
| Yes | 20 (8.3) | 74 (54.4) | 156 (80.8) | 51 (42.5) | 301 (43.6) |
| No | 220 (91.3) | 60 (44.1) | 36 (18.7) | 68 (56.7) | 384 (55.7) |
| Not recorded | 1 (0.4) | 2 (1.5) | 1 (0.5) | 1 (0.8) | 5 (0.7) |

| Characteristic | Upfront, stage I (n = 241) | Upfront, stage II (n = 136) | Upfront, stage III (n = 193) | Neoadjuvant, ypStage I to III (n = 120) | All (n = 690) |
|---|---|---|---|---|---|
| **HER2 status** | | | | | |
| Positive | 8 (3.3) | 9 (6.6) | 9 (4.7) | 7 (5.8) | 33 (4.8) |
| Negative | 186 (77.2) | 104 (76.5) | 152 (78.8) | 92 (76.7) | 534 (77.4) |
| Not tested or not recorded | 47 (19.5) | 23 (16.9) | 32 (16.6) | 21 (17.5) | 123 (17.8) |
| **HER2 immunohistochemistry** | | | | | |
| 0 | 125 (51.9) | 59 (43.4) | 84 (43.5) | 49 (40.8) | 317 (45.9) |
| 1+ | 61 (25.3) | 43 (31.6) | 62 (32.1) | 38 (31.7) | 204 (29.6) |
| 2+ | 30 (12.4) | 22 (16.2) | 33 (17.1) | 19 (15.8) | 104 (15.1) |
| 3+ | 8 (3.3) | 7 (5.1) | 5 (2.6) | 7 (5.8) | 27 (3.9) |
| Not tested or not recorded | 17 (7.1) | 5 (3.7) | 9 (4.7) | 7 (5.8) | 38 (5.5) |
| **Mismatch repair (immunohistochemistry)** | | | | | |
| Deficient (dMMR) | 10 (4.1) | 17 (12.5) | 17 (8.8) | 3 (2.5) | 47 (6.8) |
| Proficient (pMMR) | 217 (90.0) | 117 (86.0) | 169 (87.6) | 111 (92.5) | 614 (89.0) |
| Not tested or not recorded | 14 (5.8) | 2 (1.5) | 7 (3.6) | 6 (5.0) | 29 (4.2) |
| **Epstein-Barr virus (EBER)** | | | | | |
| Positive | 29 (12.0) | 11 (8.1) | 17 (8.8) | 9 (7.5) | 66 (9.6) |
| Negative | 188 (78.0) | 120 (88.2) | 165 (85.5) | 99 (82.5) | 572 (82.9) |
| Not tested or not recorded | 24 (10.0) | 5 (3.7) | 11 (5.7) | 12 (10.0) | 52 (7.5) |
| **Ki-67 index, %** | | | | | |
| Tested | 223 (92.5) | 134 (98.5) | 187 (96.9) | 111 (92.5) | 655 (94.9) |
| Median (IQR) | 70 (60 to 80) | 70 (60 to 80) | 70 (60 to 80) | 70 (30 to 80) | 70 (50 to 80) |
| **PD-L1 combined positive score** | | | | | |
| Tested | 0 (0.0) | 3 (2.2) | 5 (2.6) | 8 (6.7) | 16 (2.3) |
| CPS 1 or more, of tested | 0 tested | 0 of 3 | 2 of 5 | 3 of 8 | 5 of 16 |

Values are n (% of all patients in the group) unless stated, so the "not tested or not recorded" rows show testing completeness. Stage is pTNM after upfront surgery and ypTNM after neoadjuvant chemotherapy. HER2 status is as recorded in the final pathology report. HER2 status is unknown in 85 patients with immunohistochemistry 2+ and no recorded in situ hybridisation result and in 38 without immunohistochemistry. Mismatch repair was assessed by immunohistochemistry; microsatellite status was recorded identically and is shown with it. Laurén type is not reported because it was recorded for only 291 patients (42.2%). IQR, interquartile range; CPS, combined positive score.

### Table S2. Follow-up completeness, surveillance testing and detected recurrence among patients followed at the treating hospital, 2019 cohort

| Measure | Upfront, stage I (n = 178) | Upfront, stage II (n = 107) | Upfront, stage III (n = 142) | Neoadjuvant, ypStage I to III (n = 107) | All (n = 534) |
|---|---|---|---|---|---|
| **Follow-up completeness** | | | | | |
| Hospital-based follow-up, months, median (IQR) | 26.5 (9.3 to 61.2) | 24.1 (6.5 to 61.4) | 20.8 (7.9 to 48.0) | 15.9 (6.1 to 37.2) | 21.2 (7.4 to 58.1) |
| Five-year survivors still seen at the hospital at 48 months or later | 75 of 172 (43.6) | 39 of 94 (41.5) | 34 of 81 (42.0) | 18 of 51 (35.3) | 166 of 398 (41.7) |
| **Patients with at least one test within five years, n (%)** | | | | | |
| Serum tumour markers | 166 (93.3) | 104 (97.2) | 129 (90.8) | 102 (95.3) | 501 (93.8) |
| Abdominal CT | 144 (80.9) | 97 (90.7) | 126 (88.7) | 98 (91.6) | 465 (87.1) |
| Chest CT | 112 (62.9) | 71 (66.4) | 98 (69.0) | 88 (82.2) | 369 (69.1) |
| Gastroscopy | 104 (58.4) | 50 (46.7) | 55 (38.7) | 31 (29.0) | 240 (44.9) |
| Colonoscopy | 55 (30.9) | 18 (16.8) | 20 (14.1) | 14 (13.1) | 107 (20.0) |
| PET-CT | 6 (3.4) | 12 (11.2) | 26 (18.3) | 25 (23.4) | 69 (12.9) |
| **Tests per patient-year alive, year 1** | | | | | |
| Serum tumour markers | 1.99 | 3.29 | 4.06 | 5.55 | 3.50 |
| Abdominal CT | 1.10 | 1.44 | 1.72 | 2.24 | 1.55 |
| Chest CT | 0.45 | 0.90 | 1.06 | 1.81 | 0.97 |
| Gastroscopy | 0.35 | 0.22 | 0.22 | 0.16 | 0.25 |
| Colonoscopy | 0.08 | 0.04 | 0.04 | 0.12 | 0.07 |
| PET-CT | 0.01 | 0.03 | 0.07 | 0.09 | 0.04 |
| **Tests per patient-year alive, year 2** | | | | | |
| Serum tumour markers | 0.92 | 1.41 | 1.88 | 2.90 | 1.62 |
| Abdominal CT | 0.72 | 0.89 | 0.98 | 1.23 | 0.92 |
| Chest CT | 0.28 | 0.52 | 0.58 | 0.95 | 0.53 |
| Gastroscopy | 0.42 | 0.27 | 0.32 | 0.19 | 0.32 |
| Colonoscopy | 0.10 | 0.05 | 0.05 | 0.02 | 0.06 |
| PET-CT | 0.02 | 0.03 | 0.08 | 0.17 | 0.06 |
| **Tests per patient-year alive, years 3 to 5** | | | | | |
| Serum tumour markers | 0.48 | 0.79 | 0.93 | 1.10 | 0.74 |
| Abdominal CT | 0.43 | 0.58 | 0.62 | 0.73 | 0.55 |
| Chest CT | 0.37 | 0.51 | 0.49 | 0.68 | 0.47 |
| Gastroscopy | 0.30 | 0.20 | 0.21 | 0.17 | 0.24 |
| Colonoscopy | 0.13 | 0.05 | 0.08 | 0.05 | 0.09 |
| PET-CT | 0.01 | 0.02 | 0.04 | 0.05 | 0.03 |

| Measure | Upfront, stage I (n = 178) | Upfront, stage II (n = 107) | Upfront, stage III (n = 142) | Neoadjuvant, ypStage I to III (n = 107) | All (n = 534) |
|---|---|---|---|---|---|
| **Recurrence detected during hospital follow-up** | | | | | |
| Patients with detected recurrence | 7 (3.9) | 10 (9.3) | 34 (23.9) | 36 (33.6) | 87 (16.3) |
| Months to detection, median (IQR) | 58.7 (10.9 to 63.3) | 41.0 (20.6 to 48.1) | 18.1 (11.3 to 37.8) | 15.3 (7.0 to 23.6) | 17.8 (9.6 to 38.9) |
| First detected by abdominal CT | 4 | 3 | 15 | 11 | 33 |
| First detected by PET-CT | 2 | 3 | 15 | 16 | 36 |
| First detected by chest CT | 0 | 0 | 1 | 3 | 4 |
| First detected by gastroscopy | 0 | 1 | 0 | 1 | 2 |
| First detected by other test | 1 | 3 | 3 | 5 | 12 |
| Deaths without a recorded recurrence | 6 of 9 | 7 of 13 | 39 of 64 | 33 of 63 | 85 of 149 |

Restricted to the 534 patients (77.4%) whose postoperative follow-up took place at Ruijin Hospital; vital status was complete for all 690 patients regardless of where follow-up occurred. Tests per patient-year are the number of tests in the period divided by person-years alive in that period. Recurrence is the first imaging or endoscopic detection of recurrence or metastasis recorded at Ruijin Hospital; recurrences diagnosed elsewhere are not captured, so these counts are a lower bound. Hospital-based follow-up excludes two patients, one without a recorded last hospital visit and one whose recorded last visit precedes surgery. A patient is counted as tested within five years if at least one test was recorded in the first 60 months. Where two tests detected recurrence on the same day, the first-listed test is credited. One patient with recurrence recorded on chest CT but no recorded date is not counted. In one patient the cumulative tumour marker count decreased between periods; negative increments were set to zero.
